# MatchMiner: An open source platform for cancer precision medicine

**DOI:** 10.1101/2022.02.02.22270186

**Authors:** Harry Klein, Tali Mazor, Ethan Siegel, Pavel Trukhanov, Andrea Ovalle, Catherine Del Vecchio Fitz, Zachary Zwiesler, Priti Kumari, Bernd Van Der Veen, Eric Marriott, Joyce Yu, Adem Albayrak, Susan Barry, Rachel B. Keller, Laura E. MacConaill, Neal Lindeman, Bruce E. Johnson, Barrett J. Rollins, Khanh T. Do, Brian Beardslee, Geoffrey Shapiro, Suzanne Hector-Barry, John Methot, Lynette Sholl, James Lindsay, Michael J. Hassett, Ethan Cerami

## Abstract

The systematic deployment of next generation sequencing means patient tumors can be genomically profiled and specific genetic alterations can be targeted with precision medicine (PM) drugs. More therapeutic clinical trials are needed to test new PM drugs to advance precision medicine, however, the availability of comprehensive patient sequencing data coupled with complex clinical trial eligibility has made it challenging to match patients to PM trials. To facilitate enrollment onto PM trials, we developed MatchMiner. MatchMiner is an open-source platform to computationally match genomically profiled cancer patients to PM trials. Here, we describe MatchMiner’s capabilities, outline its deployment at Dana-Farber Cancer Institute (DFCI), and characterize its impact on PM trial enrollment. MatchMiner’s two primary goals are to (1) facilitate PM trial options for all patients, and (2) accelerate trial enrollment onto PM trials. MatchMiner has 3 main modes of use: (1) patient-centric, where a clinician looks up trial options for an individual patient, (2) trial-centric, where a trial team identifies candidate patients for their trial by setting up a filter, and (3) trial search, where a clinician can find trial options for patients that have external genomic reports. From the time MatchMiner was first deployed at DFCI in March 2016 through March 2021, we curated 354 PM trials containing a broad range of genomic and clinical eligibility criteria and MatchMiner facilitated 166 trial consents (MatchMiner consents, MMC) for 159 patients. To quantify MatchMiner’s impact on trial consent, we retrospectively measured time from genomic sequencing report date to trial consent date for the 166 MMC compared to trial consents not facilitated by MatchMiner (non-MMC). We found MMC consented to trials 55 days (22%) earlier than non-MMC. MatchMiner has enabled our clinicians to match patients to PM trials and accelerated the trial enrollment decision making process.

## Introduction

Genomic profiling of patient tumors has become an integral part of cancer care as new druggable targets are discovered and precision medicine (PM) treatments gain widespread use ^1–5^. Drugs targeting specific alterations such as *EGFR* mutations, *BCR-ABL* fusions, and *BRAF* mutations have provided great clinical benefit ^6–8^. To continue to advance the state of cancer therapy, more patients need to participate in clinical trials to test new PM drugs. Despite the more common use of genomic profiling, as few as 10-15% of patients with profiled actionable mutations participated in genotype-driven trials ^9–13^. Low PM trial participation can be caused by several factors including low clinician awareness of eligible trials, patient performance status, and patient attitudes and financial concerns ^14–16^.

Another barrier for trial participation is matching patient genomic data to PM trial eligibility criteria. Without advanced trial matching systems, individual oncologists must track hundreds of active clinical trials, only a few of which may be relevant for any given patient ^17,18^. In addition, PM trials are often basket trials that enroll patients across histologies with similar genomic changes, making recruiting patients across multiple departments a laborious process ^19–21^. Thus, an informatics system would be useful to integrate both genomic data and clinical trial eligibility criteria to support PM trial enrollment.

At Dana-Farber Cancer Institute (DFCI) over 40,000 patient tumor samples have been genomically profiled. Patient tumors are sequenced with two next generation sequencing (NGS) panels: (1) OncoPanel, which identifies mutations, copy number alterations, structural variants, and mutational signatures in ∼450 cancer relevant genes ^22^, and (2) Rapid Heme Panel, which identifies mutations and copy number alterations in 88 genes relevant in hematological malignancies ^23^. DFCI uses OnCore for its clinical trial management system and Epic for patient electronic health records (EHR). To integrate these DFCI systems into a single platform for PM trial matching and viewing patient and trial data, we developed MatchMiner, an open-source software platform for matching cancer patients to PM trials.

The goals of this manuscript are to (1) outline the core functionalities of MatchMiner; (2) describe the capabilities of Clinical Trial Markup Language (CTML) and how it is used to encode PM trial eligibility criteria for MatchMiner; (3) explain how MatchMiner is used at DFCI; (4) characterize the PM trials that have been curated into MatchMiner; and (5) assess the clinical utility of MatchMiner by a) characterizing the PM trial consents that have occurred because of MatchMiner and b) determining the impact of MatchMiner on the speed with which patients consent to PM trials.

## Results

### MatchMiner capabilities

We designed MatchMiner to provide patient matches to PM trials. MatchMiner trial matching is performed via the MatchEngine, an algorithm that computes trial matches based on patient genomic and clinical data and PM trial eligibility criteria (Fig. 1a). The MatchEngine accepts many different data inputs for patient-trial matching and therefore is adaptable to data available at any institution. Data inputs can include: (1) patient-specific genomic sequencing data, including mutations, copy number alterations (CNA), structural variants, tumor mutational burden and mutational signatures including mismatch repair deficiency, tobacco, and UV light, patient-specific clinical data, including primary cancer type, gender, age, and vital status, and trial criteria including genomic eligibility, cancer type, age, and accrual status. MatchMiner can accept a range of genomic specificity in trial matching, ranging from any mutation in a gene to specific amino acid changes. In addition to trial matching, patient genomic reports and PM trial information are viewable in a user-friendly format, providing context to trial matching results and acting as a resource for clinicians to view current PM trials. For data integrity and security standards, MatchMiner meets HIPAA requirements when deployed within an institutional firewall.

**Figure 1.**
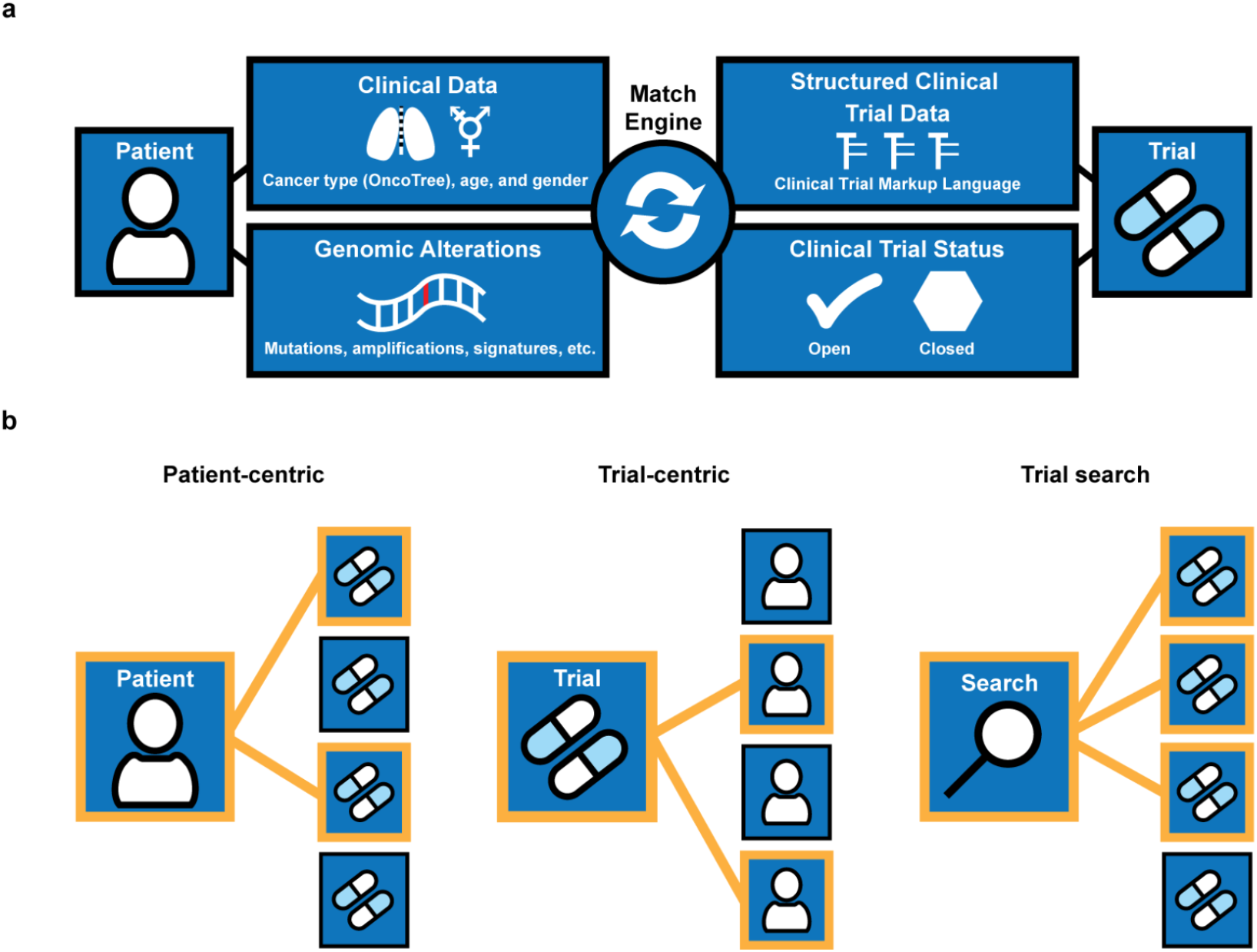
MatchMiner overview of data flow and modes of use. (a) Data inputs from patients and trials are utilized by the MatchEngine to match patients to trials. (b) Shown are the 3 modes of matching patients to trials: patient-centric, trial-centric, and trial search. Orange lines indicate patient-trial matches.

MatchMiner has several modes of clinical use: (1) patient-centric, where clinicians look up patients to view all trial matches for that particular patient, (2) trial-centric, where clinical trial teams identify patients for their particular PM trials of interest, and (3) trial search, where clinicians manually enter search criteria of interest to identify available trials based on external genomic reports (Fig. 1b). In general, MatchMiner serves as a pre-screening tool since not all trial eligibility criteria are included in the matching process and there is no consideration about a patient’s readiness to participate in a trial.

In patient-centric mode, a trial match summary page displays all potential NGS-based trial matches and highlights the genomic alteration(s) responsible for each trial match (Supplementary Fig. 1). While matches are shown per trial, MatchMiner actually matches patients to specific arms of a trial, providing additional precision in the trial match results. Trial matches are ranked according to variant actionability and can be filtered according to genomic targets or other trial features. A clinician can also (1) view additional trial details (e.g., number of arms, status of each arm, individual trial arm eligibility criteria) to determine whether a trial is suitable for their patient; (2) contact the trial investigator through an embedded email icon; and (3) access the patient’s full NGS report. Trial matches are updated nightly with matches to currently enrolling arms of open trials, making the patient-centric mode an up-to-date resource for trial options.

In trial-centric mode, a clinical trial team sets up a filter based on the genomic and clinical features of interest for a particular trial (Supplementary Fig. 2a). Filters match against all living patients in the system, and matches (i.e., all candidate patients for their trial) are viewed on the “Matches” page (Supplementary Fig. 2b). This page displays select clinical and genomic features of each trial match and provides a link to email a patient’s physician to help determine their eligibility and interest in a PM trial. As patient eligibility is reviewed, clinicians can place patients in “bins” according to their eligibility status. Filter matches are updated nightly, and clinical trial team members receive an email when new matches are identified for their filters. Thus, trial-centric mode is a method to identify new candidate patients for a trial in real-time.

In trial search mode, users can explore all the curated trials that are used to generate patient-centric matches (Supplementary Fig. 3). The trial search page can be used to find matches for patients who have had external NGS (i.e., their NGS results are not available within MatchMiner). Genomic and clinical features from the external report are inputted manually and all available trials are searched. A faceted search interface allows for further filtering based on multiple criteria, including genomic changes, cancer type, trial status, and trial phase. The trial search page is also a convenient place for clinicians to view all the latest available PM trials at their institution, including arm status and trial eligibility information. For example, all trials targeting BRAF can be viewed, or specific drugs of interest can be searched. Trial search mode helps match patients with external genomic sequencing data to PM trials and acts as a resource for clinicians to view PM trial information.

To ensure results provided in each clinical mode of use are timely, MatchMiner requires daily data updates. Updated genomic data (e.g. from an institutional enterprise data warehouse), patient vital status (e.g. from Epic or other EHR) and trial accrual status at both the trial and arm level (e.g. from OnCore or other trial management system) are ingested daily. Following the daily data update, the MatchEngine runs to compute updated matches for all living patients to all open arms of open trials, as well as to all filters. As a result of these daily updates, MatchMiner is able to provide clinicians with accurate and timely trial matches.

### Clinical trial markup language (CTML)

Protocols for PM trials are often lengthy documents that contain genomic and clinical eligibility criteria in an unstructured format, making it difficult to extract information for patient-trial matching ^24^. To structure genomic eligibility data for trial matching, we developed clinical trial markup language (CTML). CTML is a human-readable markup language that allows users to structure clinical trial details including clinical and genomic eligibility. While CTML may be used standalone as a means of encoding trial eligibility, its main utility is as part of a trial matching system. Trial eligibility criteria can be translated into CTML documents via a text editor. Alternatively, an open-source curation platform allows a user to pull in criteria from clinicaltrials.gov^25^ – this can be helpful for extracting basic trial information, but specific genomic and clinical eligibility may need to be entered manually.

CTML supports a wide range of clinical trial information and is easily extended to include new criteria. Eligibility criteria, including genomic, clinical, and demographic criteria, are encoded with nested Boolean logic. In trial protocols, eligibility criteria are typically written in a numbered list with subcriteria (Fig. 2a). CTML files are structured similarly to clinical trial eligibility criteria but have defined core elements. They have an intrinsic tree-like structure anchored by core trial details and extended by arms and dose levels (Fig. 2b). Each arm and dose level component of a trial may have its own distinct eligibility criteria, which is encoded in the CTML document. The flexibility to encode eligibility criteria, as well as accrual status, at any arm or dose enables CTML to accurately capture complex trial structures and results in more precise trial matches.

**Figure 2.**
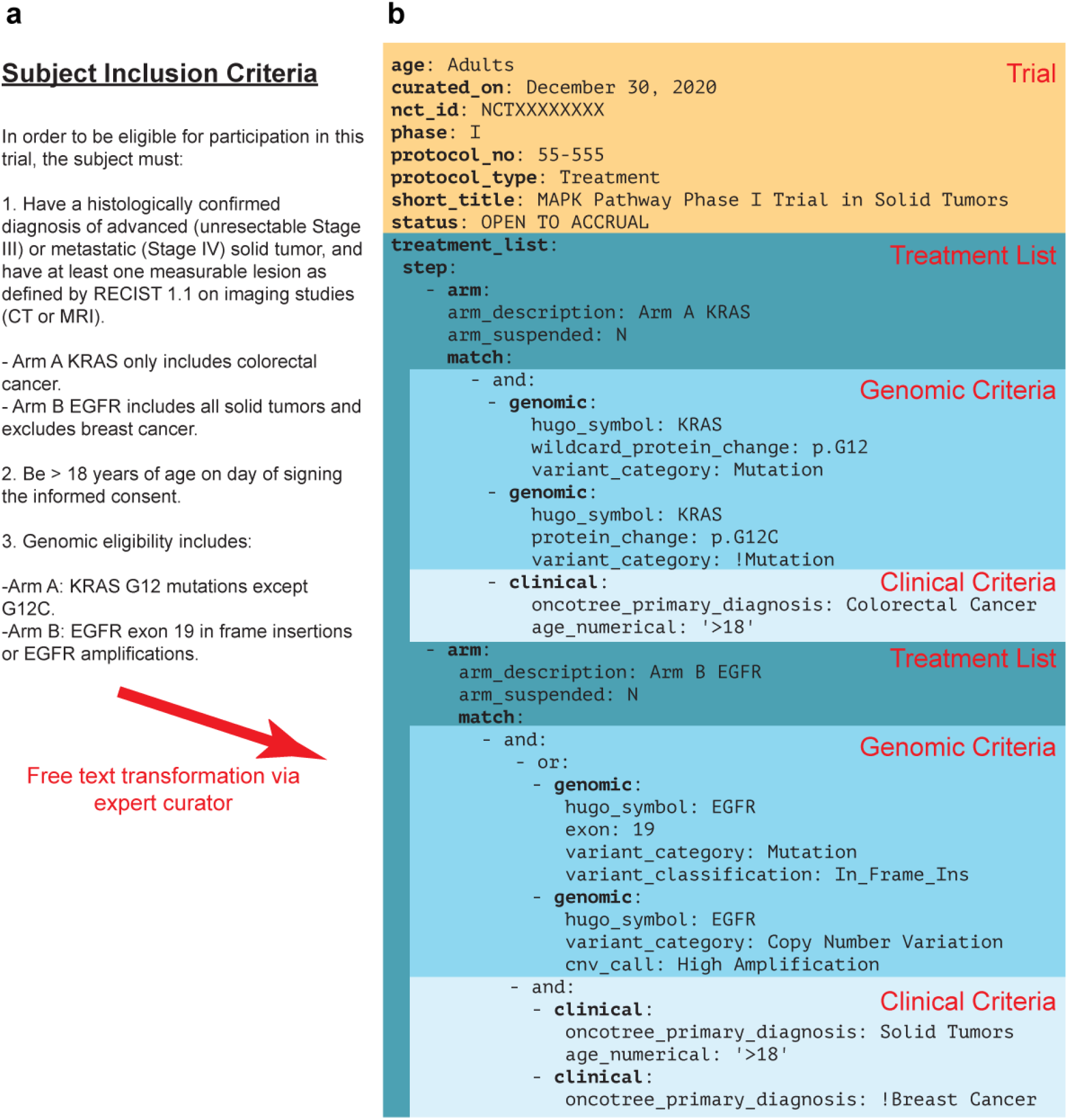
Clinical trial markup language (CTML) provides a structured data format for PM trial eligibility criteria. (a) Free text subject eligibility criteria from an example PM trial protocol. Arm A is enrolling patients with colorectal cancer and any KRAS G12 mutation except G12C. Arm B is enrolling patients with EGFR altered (specifically EGFR exon 19 insertions or EGFR amplifications) solid tumors except breast cancer. (b) Trial details transformed into CTML, with curated information related to basic trial metadata (orange) and the treatment arms (dark blue) containing specific genomic (turquoise) and clinical (light blue) match criteria. Arm A and B exclusions are annotated using exclamation points.

CTML has five core components:

- **Trial**: Basic metadata, such as short and long trial titles, the national clinical trial (NCT) purpose and identifier from the public registry of trials at clinicaltrials.gov, contact information, and study phase.
- **Treatment**: Steps, arms, doses, and expansion cohorts of a clinical trial, which form a tree-like structure for complex trials. Nodes of this tree have match criteria which contain genomic and clinical eligibility information.
- **Match**: Contains Boolean logic specifying genomic and clinical criteria. Additional match components are referenced via logical operators (and/or/not).
- **Clinical**: A Boolean clause that contains specific clinical criteria. Currently supported fields are age, gender, and cancer type according to the OncoTree ontology ^26^.
- **Genomic**: A Boolean clause that contains specific genomic criteria. Currently, supported alterations include mutations, which can be restricted to specific types of mutations, exon locations or protein changes, gene-level copy number alterations, structural variants and mutational signatures.

From a trial matching perspective, the most important part of any CTML document is the match clause. All match clauses must be enclosed within an “and” or “or” clause which contains eligibility attributes. “And” and “or” clauses may be nested within each other and there is no limit to the depth or breadth of a single match clause. Any clinical or genomic criteria can be marked as an exclusion criteria by prefacing the relevant criteria with “!” (Fig. 2b). For matching purposes, the eligibility criteria within CTML match clauses must reflect the available patient data. For example, if a patient’s date of birth is unavailable, then the trial curations should not include an age restriction. Thus, the match clause must consist of available data and be accurate for successful trial matching.

### MatchMiner use at Dana-Farber Cancer Institute

Since the launch of MatchMiner at DFCI, we have tailored the platform according to institutional workflows and available NGS data. For ease of use in the clinic, we integrated MatchMiner patient-centric and trial search modes into Epic. This allows clinicians to review trial matches directly alongside the clinical data available in the electronic medical record to further aid decision making (Supplementary Fig. 4). When viewing a patient’s chart in Epic, a clinician can open a tab to view MatchMiner trial matches for that patient or search through all MatchMiner trials. To leverage the evolution of available NGS data at DFCI, MatchMiner has added support for additional assays and data types. Currently, MatchMiner supports trial matching based on OncoPanel data in both patient-centric and trial-centric modes^22^, while Rapid Heme Panel data is available within trial-centric mode^27^.

MatchMiner has been integrated into the specific workflows of various clinical groups. A collaboration with the Center for Cancer Therapeutic Innovation (CCTI) resulted in a MatchMiner tumor review board process, where a thorough assessment of potential trial-centric matches was performed. Each week, matches for several CCTI trials were filtered based on additional requirements, such as an upcoming appointment, and then manually reviewed by the MatchMiner team for evidence of progression in radiology scan text impressions. The resulting list of patients was reviewed with CCTI staff, and patients deemed ‘trial ready’ were flagged and their treating physician contacted. In a separate collaboration with the Gastrointestinal Cancer Center (GCC), we developed GI-TARGET, a program which integrates the patient-centric mode of MatchMiner with additional molecularly driven therapy suggestions for holistic review by a team of experts, resulting in patient-specific treatment suggestions. GI-TARGET continues to evolve and be an essential component of the GCC.

MatchMiner users span nearly all disease groups at DFCI, including Sarcoma, Breast, Thoracic, and Pediatric. For some trial-centric users, the MatchMiner team supplements the trial-centric mode by sending spreadsheets of patient-trial matches on a weekly or monthly basis. The MatchMiner team also supports the monthly DFCI pan-cancer molecular tumor board where we frequently review all available MatchMiner trial matches for specific patient cases. Thus, MatchMiner has been adapted for the specific data available at DFCI and continues to be integrated into departmental workflows for PM trial recruitment.

### Features of PM trials in MatchMiner

At DFCI, PM trials are added to MatchMiner after a structured review process. Trial protocol documents for newly opened trials are reviewed weekly and selected if they contain genomic eligibility criteria. Selected protocols are then curated into CTML and reviewed by a second team member prior to uploading into MatchMiner. CTML documents are updated whenever a new arm is added to a trial or an existing arm is removed. Trial protocol documents are also systematically reviewed every 3 months to capture any changes in eligibility from study amendments. As of March 2021, we have 354 PM trials curated in MatchMiner, reflecting all trials with genomic eligibility that have been open at DFCI since MatchMiner launched in 2016. These trials provide a breadth of options for patients, as 80% of NGS-sequenced patients have at least one trial option, with an average of 6 trial options for each patient.

To explore the landscape of PM trials available at DFCI, we quantified genomic and cancer type inclusions in the 354 trials curated in MatchMiner. A total of 222 genes, 7 mutational signatures, and 59 specific cancer types (OncoTree metatypes, 91% of 65 total metatypes) were represented ^26^. In addition, general criteria for all solid or all liquid cancer types were also commonly included as trial eligibility criteria. The gene most frequently included as an inclusion criterion across all trials was *BRAF* (n=46 trials), followed by *EGFR* (n=42 trials), and *KRAS* (n=38 trials) (Fig. 3a), reflecting the many therapies in PM trials for these oncogenic drivers ^28–30^. Other genes frequently leading to PM trial eligibility included *IDH1* (n=24) and *IDH2* (n=20), *NTRK* (n=15), *ALK* (n=27), and *ROS1* (n=12), and homologous recombination repair (HR) genes such as *BRCA1*/*BRCA2* (n=35) and *PALB2* (n=20). The majority of trials included 1-3 unique genes in their eligibility criteria (n=286, 81%) while a small proportion of trials had 4 or more unique genes (n=68, 19%) (Fig. 3b). For cancer types, trials most commonly enrolled all solid tumors (n=123), non-small cell lung cancer (n=97), and breast cancer (n=50) (Fig. 3c), consistent with the abundance of Phase I trials in solid tumors and targeted therapies for these cancer types ^1,31,32^. To explore the diversity of included cancer types outside of trials with broad all solid/liquid eligibility, we next examined how many specific cancer types are included for each trial. Excluding trials with all solid/liquid eligibility, the majority of trials enroll one specific cancer type (n=185, 81%) and few trials enroll 2 or more specific cancer types (n=44, 19%) (Fig. 3d).

**Figure 3.**
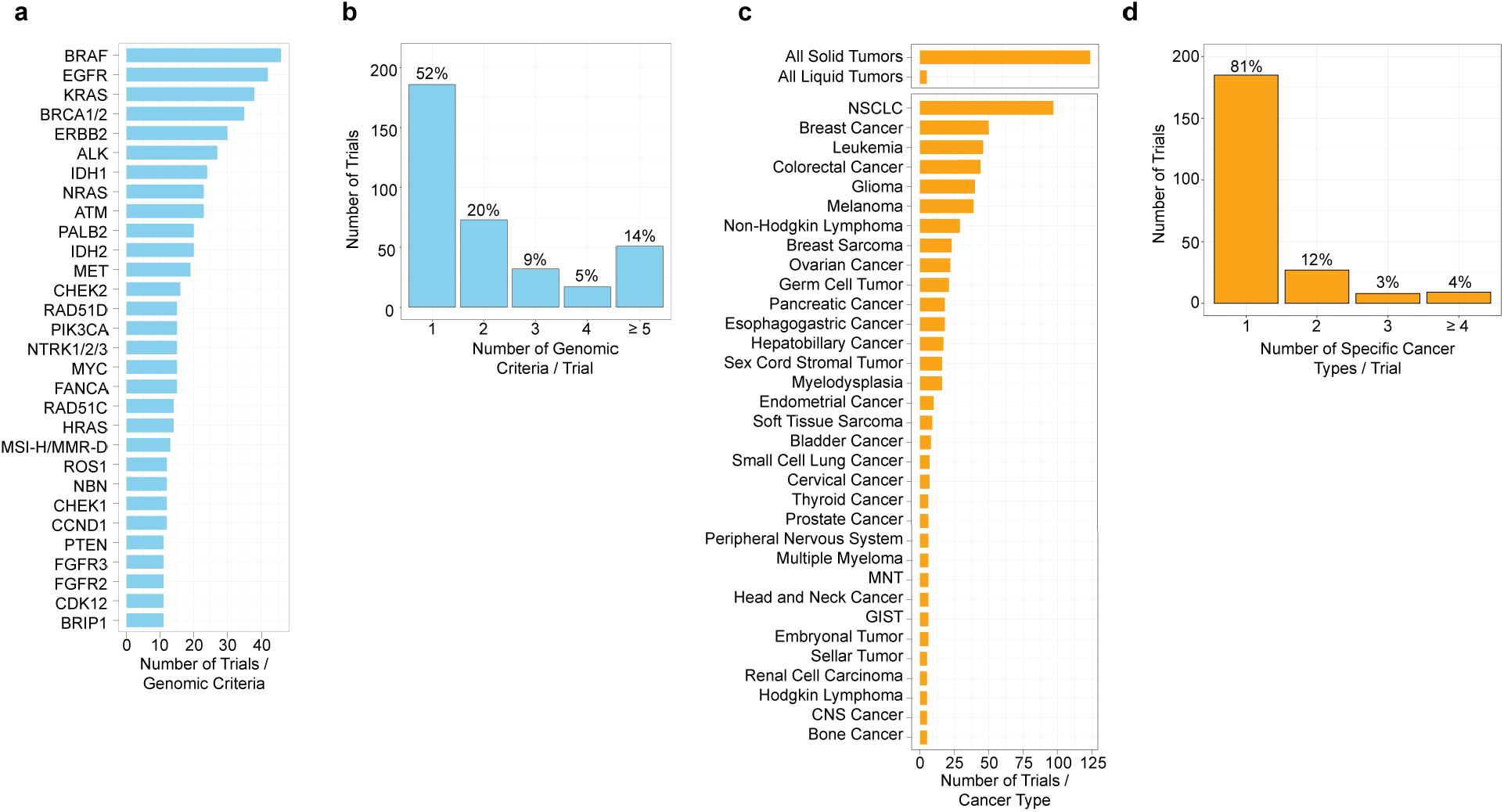
The landscape of genes and cancer types in DFCI precision medicine trials. (a, b) Number of trials in MatchMiner for select genomic criteria (≥ 10 trials) and number of trials that have 1, 2, 3, 4, and ≥ 5 genomic criteria. (c, d) Number of trials in MatchMiner for select cancer types (> 4 trials) and number of trials that have 1, 2, 3, and ≥ 4 specific cancer types. NSCLC = non-small cell lung cancer, GIST = gastrointestinal stromal tumor. MNT = Miscellaneous Neuroepithelial Tumor. Trials with eligibility for all solid or all liquid tumors were not included in (d).

In addition to quantifying genes and cancer types, we also examined the distribution of trial phases and the disease centers running each trial. Phase I (38%) and Phase II trials (31%) were the most common, followed by Phase I / II (17%), Phase III (11%), and Phase II / III (0.8%) trials (Supplementary Table 1). The higher proportion of earlier phase trials is consistent with the large number of novel PM drugs emerging, and the fact that most drugs do not progress to later phase trials ^33^. Most trials (23%) were run out of the CCTI followed by thoracic oncology (17%) and pediatric oncology (11%). Thus, MatchMiner at DFCI has mostly Phase I and II trials, involving a range of genomic criteria and cancer types.

### Impact of MatchMiner on PM trial consent

We analyzed PM trial enrollment data to determine whether MatchMiner led to earlier identification of trials among NGS-sequenced patients. We have identified 166 MatchMiner patient consents, derived from 159 patients (7 patients consented to multiple trials) and 65 trials. These 166 patient consents were attributed to MatchMiner (i.e., considered MatchMiner consents [MMC]), because MatchMiner identified the potential match and the provider or trial team viewed the match in MatchMiner prior to the patient consenting to the PM trial. The average age of MMC patients is 60 years old (min=8, max=86) with most patients between 50-64 years old (n=70, 44%) (Supplementary Table 2). 67% of patients are female (n=106) and 33% are male (n=53). Most patients are white (n=137, 86%), followed by African American (n=7, 4%), Asian (n=6, 4%), and other/unknown (n=6, 4%). The socio-demographic features of the MMC cohort are similar to the total DFCI patient population.

To further assess the impact of MatchMiner, we compared the time to consent (i.e., time from NGS sequencing to PM trial consent) for the 166-patient MMC cohort to a ‘control’ population. The control population was composed of patients who also had NGS sequencing and had consented to one of the 65 trials for which there was a MatchMiner-linked consent, but for which there was no record that their matches had been viewed in MatchMiner.

As outlined above, the primary outcome was the number of days from when an OncoPanel report was uploaded into MatchMiner (i.e., the MatchMiner ingestion date) to when the patient consented to the PM study. We chose this outcome because clinicians only have the potential to view trial matches after an OncoPanel report is added to MatchMiner. We found the time to consent for the MMC was 55 days faster than for the non-MMC cohort (195 days [IQR=85-341 days] vs 250 days [IQR=99-491 days], P=0.004, Fig. 4b). An analysis of the distribution of time-to-consent data for the MMC and non-MMC cohorts found no evidence that outliers were skewing our data. Thus, MatchMiner may provide clinical impact by accelerating time to consent for these PM trials.

**Figure 4.**
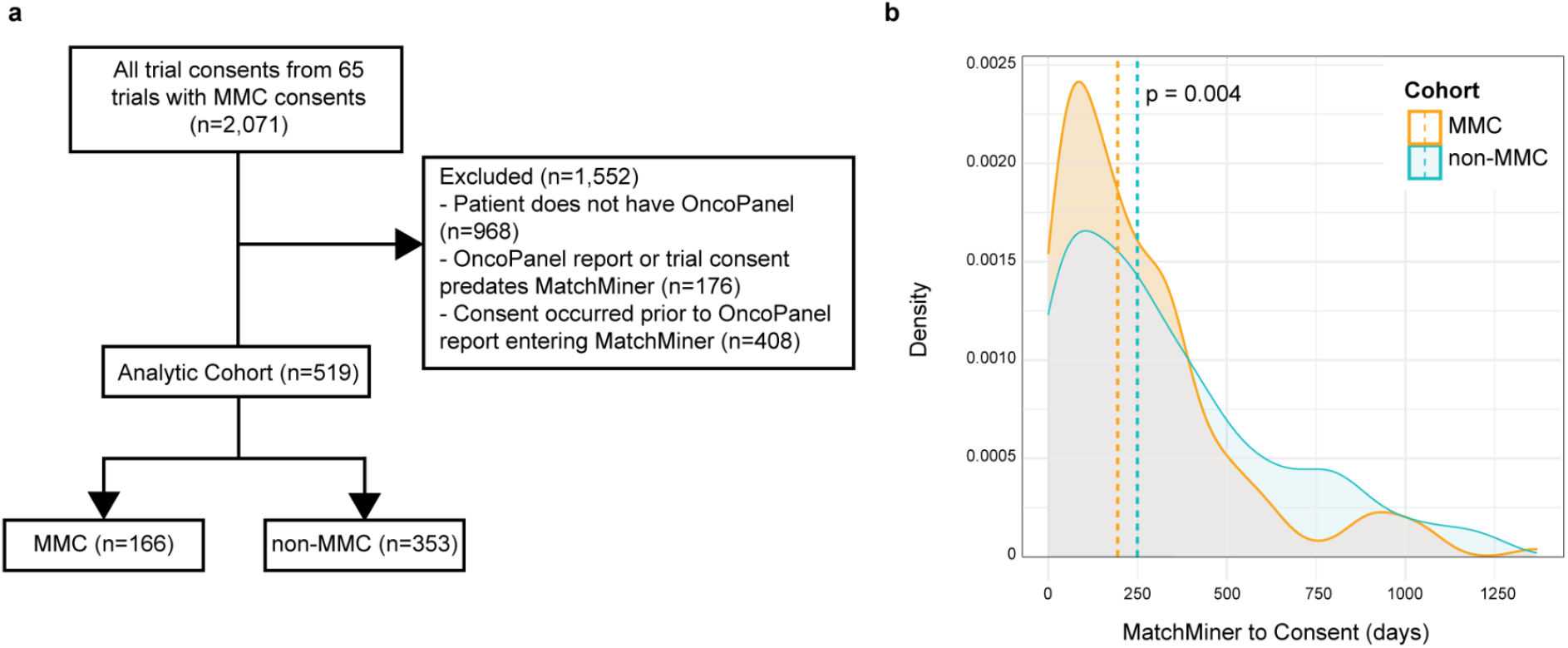
MatchMiner decreases time to consent for precision medicine trials. (a) 2,071 consents from 65 trials in MatchMiner were filtered to generate comparison consents for MMC. 1,552 consents were excluded due to the patient not having an OncoPanel report or only having a failed OncoPanel report, the OncoPanel report or trial consent date were prior to the launch of MatchMiner, or the trial consent date preceded the ingestion of the OncoPanel report in MatchMiner. The remaining 519 enrollments were divided into MMC and non-MMC. (b) Density plot of time period from MatchMiner ingestion date to consent date. MMCs had a median of 195 days (IQR = 85, 341 days) compared to non-MMCs with a median of 250 days (IQR = 99, 491 days). Medians compared with a Wilcoxon rank-sum test.

## Discussion

Here, we described MatchMiner, an open-source software platform for matching cancer patients to PM trials. Previous informatics platforms have been developed to match patients to PM trials including academic cancer center solutions ^17,34^ and commercial solutions from companies such as Foundation Medicine, IBM Watson, and Syapse. However, the impact of these solutions on clinically relevant outcomes is not well characterized, many of these solutions are proprietary, and some of these solutions are not portable to other institutions. MatchMiner is a trial matching platform that solves many of these limitations and has clinical impact at DFCI.

MatchMiner decreased time to consent by 55 days, demonstrating impact for a clinically relevant outcome. Timely trial enrollment is critical for patient care in part because a major barrier for trial enrollment is poor performance status. Earlier trial identification could provide options for patients before performance status declines ^15,16^. Timely enrollment is also critical for PM trials, where it can be difficult to find genomically eligible patients. In addition, MatchMiner decreased time to PM trial enrollment regardless of the mode of use (patient-centric, trial-centric or collaboration with clinical groups).

We are aware that there are potential confounding factors because we performed a retrospective analysis. For example, we were not able to control for interdepartmental variability in the trial enrollment process. We also were not able to control for the number of enrollments for each trial and individual trial recruitment rates. Despite these limitations, a significant median difference of 55 days between MMC and non-MMC is strongly suggestive of clinical impact for MatchMiner.

In addition to MatchMiner’s clinical impact, we highlighted several key advantages that make MatchMiner suitable as a clinical tool for trial matching. (1) MatchMiner has consistent, structured genomic eligibility criteria using the CTML standard, which can be easily adapted for data inputs at other institutions. CTML allows complex eligibility criteria to be curated for more accurate matching. (2) MatchMiner gives clinicians real-time access to comprehensive structured NGS and trial status data. Real-time trial matching allows clinicians to make more accurate trial enrollment decisions. (3) With EHR integration, MatchMiner can be more easily adopted into existing clinical workflows. At DFCI, we integrated MatchMiner into Epic, allowing departments easier access to MatchMiner and more efficient cross-referencing with patient medical history. These advantages, in addition to being open-source, make MatchMiner a viable option for adoption at other institutions nationwide.

While MatchMiner has been successfully established as a trial matching tool throughout DFCI, we are also focused on continuing to grow the platform. Integrating all of MatchMiner’s features into an EHR system (e.g. Epic) is challenging, especially for an open-source platform that aims to be EHR-vendor agnostic. To address this issue, we are exploring open standards for future releases such as SMART on FHIR, a set of open specifications to integrate apps with EHRs ^35,36^. A second major challenge is integrating additional clinical eligibility criteria, such as prior therapies and laboratory values, into trial matching. This requires modifying the CTML to include additional clinical criteria, and adjustments to interoperable standards for extracting structured clinical data from EHRs. We envision proposing a formal standard for CTML via widely used collaborative standard development processes such as the Global Alliance for Genomics and Health (GA4GH) ^37^. Third, as the number of data sources integrated into MatchMiner grows, standardization and data processing workflows will become more challenging. Genomic and clinical data harmonization is important since pipelines call variants differently and cancer ontologies can differ amongst institutions ^38^. Lastly, it is difficult to determine whether a patient is ready to enroll on a trial (trial readiness) from limited clinical data. Further enrichment of patient-trial matches with radiology data, for example, could more precisely identify patients ready for trials.

In summary, MatchMiner is an open-source tool for matching patient genomic profiling to PM trials. With three modes of use, MatchMiner can be used to look up trials for individual patients or to recruit patients for a trial. MatchMiner uses CTML to structure trial eligibility criteria, and we continue to advocate for CTML as the standard for structuring trial data. MatchMiner can be used with a trial management system to show real-time trial status at the arm level. The combination of real-time trial arm status with detailed genomic eligibility down to the variant level allows MatchMiner to provide highly specific matches to PM trials. MatchMiner at DFCI has many PM trials with a range of genomic and clinical eligibility criteria. MatchMiner accelerated PM trial enrollment and future studies will aim to determine its impact within specific departments at DFCI.

## Methods

### MatchMiner technical details

MatchMiner is a two-tier web application with a mongo database serving a Python-based REST application programming interface (API) server and AngularJS 1.5 client. The MatchMiner UI is written in AngularJS and displays the user interface. The UI interfaces with the REST API and displays relevant trial match information, as well as displaying a clinical trial search interface. Institution specific ETL pipelines are used to load genomic and clinical data as well as retrieve trial data from clinical trial management systems. Authentication of users is performed via single sign-on configured through Security Assertion Markup Language (SAML).

### DFCI trial analysis

DFCI trials are run in collaboration with members of the Dana-Farber/Harvard Cancer Center (DF/HCC) consortium which includes six other Harvard-affiliated institutions, including Beth Israel Deaconess Medical Center, Boston Children’s Hospital, Brigham and Women’s Hospital, Harvard Medical School, Harvard T.H. Chan School of Public Health, and Massachusetts General Hospital. We used 354 trial json files from the DFCI MatchMiner application for our analysis. We extracted all inclusions of genes and cancer types from each of our 354 trial json files. Inclusions are defined as genes and cancer types without the “!” exclusion symbol, and genes not labeled “wild type”. Extraction of genes and cancer types was performed by recursively traversing trial CTMLs with custom scripts and summarized with R ggplot2 and gtsummary ^39,40^. All cancer types from trials were annotated with their corresponding OncoTree metatype (OncoTree version: oncotree_legacy_1.1) ^26^. Trial phases and disease center were extracted from the summary field of trial json files with Python (v. 2.7) pandas json_normalize function ^41^. Mutational signatures include tumor mutational burden, APOBEC, MSI-H/MMR-D, UVA, temozolomide, POLE, and tobacco.

### MatchMiner impact consent filtering

166 MMCs were identified through an automated system that recorded patient-centric and trial-centric page visits, followed by manual review by the MatchMiner team. For a given putative MMC, the MatchMiner team looks for evidence that a relevant clinician viewed the patient in the context of the trial prior to the patient consenting to the trial. For example, a breast oncologist viewed trial matches for one of their patients in MatchMiner and the patient then consented to one of the listed trials.

Our 166 MMC were distributed among 65 PM trials. A comparison group, non-MMC, was therefore defined as all other consents to the same 65 PM trials as the MMCs. For patients with multiple OncoPanel reports, each trial consent was paired with the closest OncoPanel report prior to the consent date. To generate the analytic cohort, we applied the following filters: (1) patient had a successful OncoPanel report, (2) the patient’s OncoPanel report and trial consent date were both after the launch of MatchMiner, and (3) the consent date is after the OncoPanel report was added to MatchMiner. After filtering, we identified 353 non-MMC to compare to our MMC.

Consent dates were extracted from DFCI’s OnCore trial registration database. For our analytic cohort, time from MatchMiner ingestion date to consent date was calculated on a DFCI HIPAA compliant server. Filtering was performed with Python (v. 2.7) pandas commands and the MatchMiner ingestion date to consent date time period was analyzed for MMC and non-MMC using a Wilcoxon rank sum test with R gtsummary ^40,41^. This study was approved by the Dana-Farber Cancer Institute/Harvard Cancer Center investigational review board (protocol #20-733), which determined that neither the physicians nor the patients needed to be consented for this retrospective study.

## Data Availability

All data produced in the present work are contained in the manuscript

## Code Availability

The MatchMiner MatchEngine, API, and UI is available in the Github repository: https://www.github.com/dfci/matchminer under a GNU Affero License. More comprehensive documentation on MatchMiner deployment and data inputs is available at https://www.matchminer.gitbook.io/matchminer/. For more information or questions about MatchMiner at DFCI please visit our website: https://www.matchminer.org.

## Acknowledgments

The authors acknowledge Elizabeth H. Williams, Chris Sander, Tamba Monrose, Michael Tung (all DFCI affiliated) for their contributions to testing and providing feedback for MatchMiner in its early stages. This work is supported by DFCI and the Fund for Innovation in Cancer Informatics (ICI).

## Author Contributions

EC and MH conceived of the project, and provided overall direction, mentoring, and manuscript editing. TM provided direction, mentoring, manuscript editing, trial curation, technical design advice and project management. HK conducted the consent analysis and contributed to writing the manuscript. AO contributed to the code, infrastructure, and trial curation. CF provided project management and trial curation. PT contributed to writing the manuscript and provided technical design advice. JY provided project management and figure design. ES, PT, ZZ, BV, EM, and JL were the primary software/devops engineers, and contributed to the code and infrastructure. PK was the data acquisition lead and contributed to the code, infrastructure and trial curation. RK contributed to trial curation, testing, and the GI-TARGET workflow. AA, SB, KD, BB, GS, and SH aided in deployment, testing, and facilitated early adoption of the platform. LM, NL, BJ, BR, JM, and LS, acted as a steering committee, provided project oversight and addressed regulatory issues. All authors read and approved the final version of the manuscript.

## Competing Interests

The authors declare no competing interests.

## Notes

### Competing Interest Statement

The authors have declared no competing interest.

### Funding Statement

This study was funded by DFCI and the Fund for Innovation in Cancer Informatics (ICI).

### Author Declarations

Institutional Review Board of Dana-Farber Cancer Institute/Harvard Cancer Center gave ethical approval for this work

### Summary of Updates

Change author name "Pavel Trukanov" to "Pavel Trukhanov".

