## Supplemental Figures and Tables for "MatchMiner: An open source platform for cancer precision medicine"

### Supplementary Figures and Tables

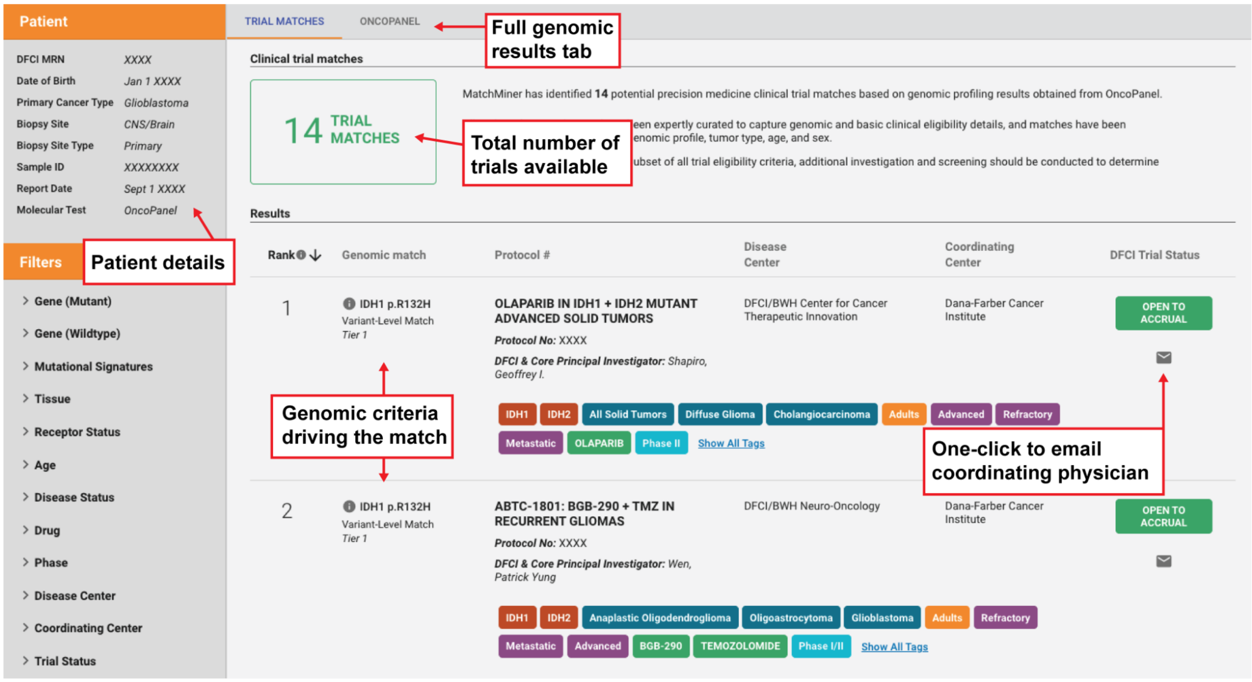


**Supplementary Figure 1. Screenshot of patient-centric trial matches page.** Shown are the available features on a simulated patient trial match page. Each row represents a patient-trial match and the genomic criteria driving the trial match is listed along with trial information. Trial matches can be filtered with faceted search according to trial genomic and clinical criteria. The coordinating physician for the trial can be emailed directly to determine slot availability and confirm patient eligibility.

a

b

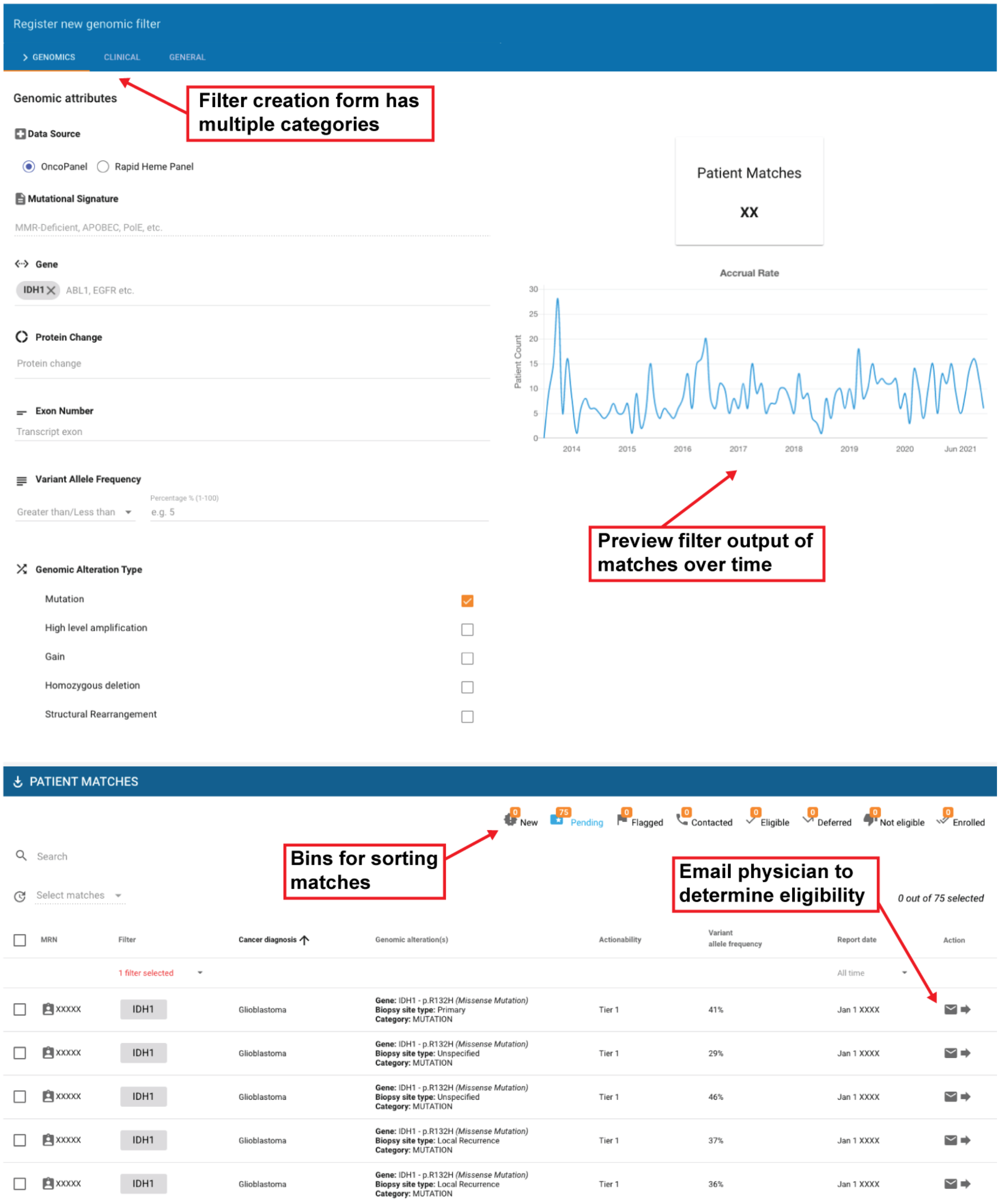


**Supplementary Figure 2. Screenshot of genomic filter creation and trial-centric matches.** (a) Genomic filter creation page with preview graph of patients with an *IDH1* mutation. Genomic criteria include mutational signatures (e.g. APOBEC), gene, protein change, alterations in a particular exon, variant allele frequency cutoff, and specific alteration types. (b) Trial matches for patients with *IDH1* mutations. Each row represents a patient-filter with fields for cancer type, genomic criteria, actionability, variant allele frequency, and report date. The patient's physician can be emailed directly to confirm patient eligibility and matches can subsequently be sorted into the provided bins.

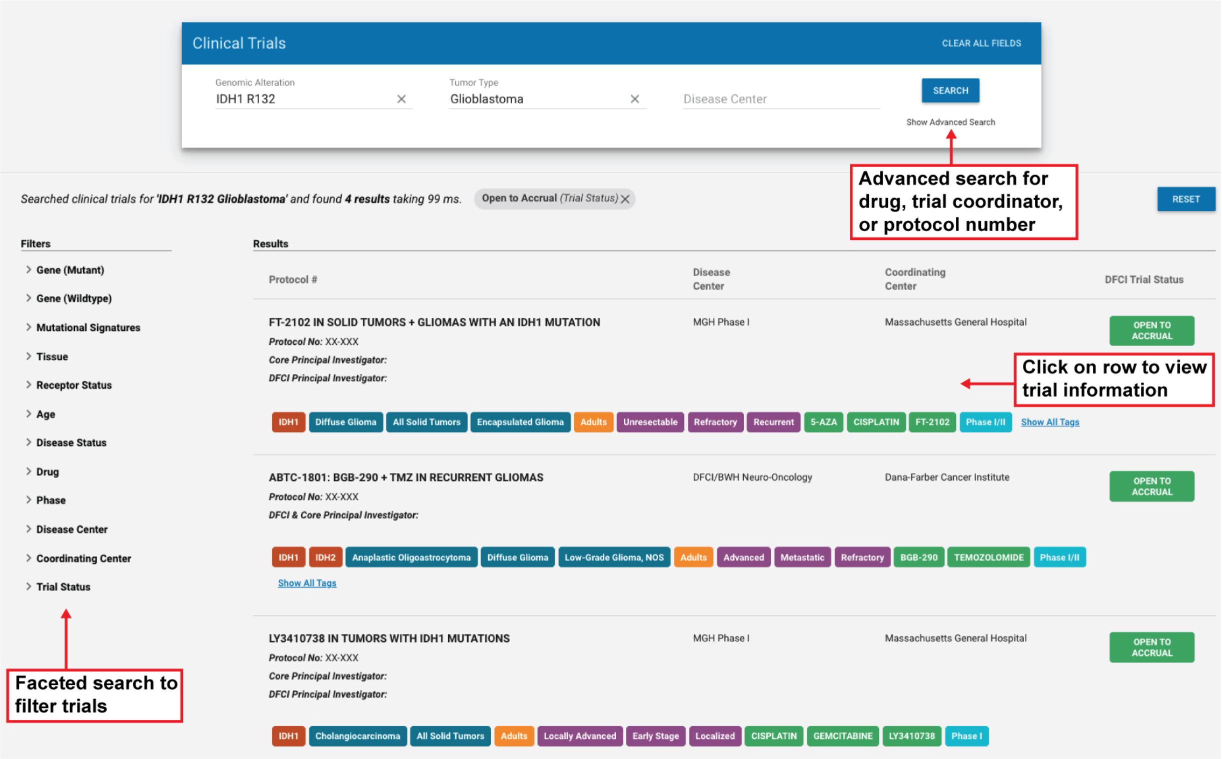


**Supplementary Figure 3. Screenshot of trial search page.** The trial search page allows the user to view details on all trials curated in MatchMiner. Trials can be searched based on specific genomic alterations, tumor type, or disease center. Advanced search allows for more specific criteria such as drug name, trial investigator, or protocol number. Trial information is viewed by clicking in the trial row.

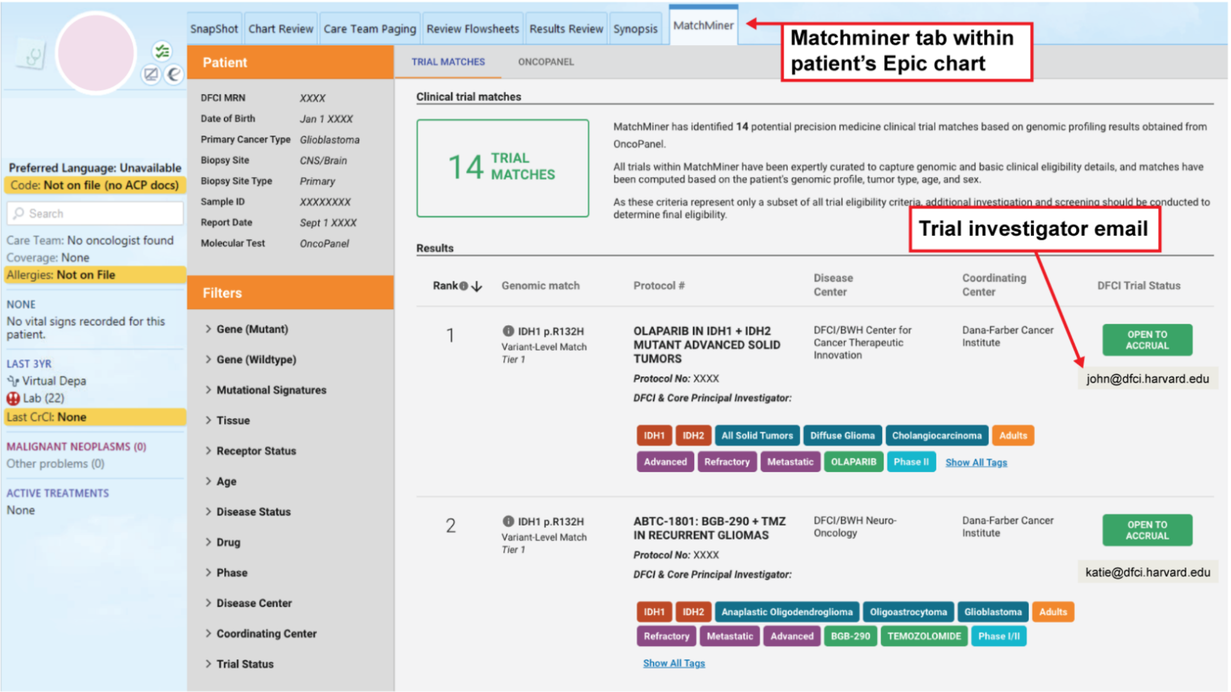


**Supplementary Figure 4. Screenshot of patient-centric trial matches in Epic.** Trial matches are shown within a patient's Epic chart. The patient's full genomic report and information on all trials in MatchMiner can also be accessed in Epic.

**Supplemental Table 1. Precision medicine trial phases and top 10 trial disease centers.**

| **Category** | **N = 354[1]** |
|---|---|
| **Trial Phase** | |
| I | 135 (38%) |
| II | 110 (31%) |
| I/II | 59 (17%) |
| III | 39 (11%) |
| Feasibility / Pilot | 8 (2.3%) |
| II/III | 3 (0.8%) |
| **Disease Center** | |
| Phase I/CCTI | 80 (22.6%) |
| Thoracic Oncology | 62 (17.1%) |

| | |
|---|---|
| Pediatric Oncology | 38 (10.6%) |
| Leukemia | 29 (8.1%) |
| Breast Oncology | 28 (8.0%) |
| Neuro-Oncology | 22 (6.2%) |
| Gastrointestinal Oncology | 20 (5.6%) |
| Melanoma | 10 (2.8%) |
| Lymphoma | 8 (2.3%) |
| Cancer Immunology | 6 (1.7%) |

[1]n (%)

**Supplemental Table 2. 159 patient demographics from the MatchMiner consents (MMC).**

| Category | MMC, N = 159[1] |
|---|---|
| **Age Group** | |
| <35 | 7 (4%) |
| 35-49 | 19 (12%) |
| 50-64 | 70 (44%) |
| 65-79 | 58 (36%) |
| 80+ | 5 (3%) |
| Mean Patient Age | 59 (52, 68)[2] |
| **Gender** | |
| Female | 106 (67%) |
| Male | 53 (33%) |

| | |
|---|---|
| **Ethnicity** | |
| Asian | 9 (6%) |
| Black or African American | 7 (4%) |
| Other/Unknown | 6 (4%) |
| White | 137 (86%) |
| **Cancer Type** | |
| Non-Small Cell Lung Cancer | 35 (22%) |
| Breast Cancer | 31 (19%) |
| Colorectal Cancer | 25 (16%) |
| Ovarian Cancer | 8 (5.0%) |
| Glioma | 2 (1.3%) |
| Pancreatic Cancer | 12 (7.5%) |
| Endometrial Cancer | 5 (3.1%) |
| Cancer of Unknown Primary | 6 (3.8%) |
| Esophagogastric Cancer | 9 (5.7%) |
| Bladder Cancer | 6 (3.8%) |
| Hepatobiliary Cancer | 2 (1.3%) |
| Gastrointestinal Stromal Tumor | 1 (0.6%) |
| Prostate Cancer | 2 (1.3%) |
| Head and Neck Cancer | 3 (1.9%) |
| Soft Tissue Sarcoma | 4 (2.5%) |
| Salivary Gland Cancer | 2 (1.3%) |
| Small Bowel Cancer | 2 (1.3%) |
| Cervical Cancer | 1 (0.6%) |

| | |
|---|---|
| Uterine Sarcoma | 1 (0.6%) |
| Skin Cancer, Non-Melanoma | 1 (0.6%) |
| Thyroid Cancer | 1 (0.6%) |

[1] n (%)

[2] Median (IQR); n (%)
